# Proteomic signatures of BMI generalize across ancestries and reveal cardiometabolic heterogeneity beyond measured BMI

**DOI:** 10.64898/2026.09.05.26362276

**Authors:** Monica Muti, Yann Ilboudo, Usama Aliyu, Chengyue Zhang, Lisa Ware, Michael Chong, Lisa K. Micklesfield, Julia H. Goedecke, Robert Ntozini, Guillaume Pare, Michael H. Cho, Michele Ramsay, Andrew P. Morris, Paul W. Franks, Abram B. Kamiza, Shane A. Norris, Omar M. E. Albagha, Matthew Moll, Tinashe Chikowore

**Affiliations:** SAMRC/Wits Ageing African Adult Research Unit (A3RU), Faculty of Health Sciences, University of the Witwatersrand, Johannesburg 2198, South Africa; Cornell University, College of Veterinary Medicine, Department of Public and Ecosystem Health, Ithaca, New York, USA; Channing Division of Network Medicine, Brigham and Women’s Hospital, Harvard Medical School.; College of Health and Life Sciences (CHLS), Hamad Bin Khalifa University (HBKU), Qatar Foundation (QF), Doha, Qatar; Diabetes Research Center, Qatar Biomedical Research Institute (QBRI), Hamad Bin Khalifa University (HBKU), Qatar Foundation (QF), P.O. Box 34110, Doha, Qatar; Population Health Research Institute, David Braley Cardiac, Vascular and Stroke Research Institute, Hamilton, Ontario, Canada; Biomedical Research and Innovation Platform, South African Medical Research Council, Cape Town, South Africa; Department of Public Health and Clinical Medicine, Umeå University, Umeå, Sweden; ZVITAMBO Institute for Maternal and Child Health Research, Harare, Zimbabwe; Sydney Brenner Institute of Molecular Bioscience, Faculty of Health Sciences, University of the Witwatersrand, Johannesburg 2193, South Africa; School of Human Development and Health, University of Southampton, UK; Department of Medical Laboratory Sciences, School of Life Sciences and Allied Health Professions, Kamuzu University of Health Sciences, Blantyre, Malawi; Arthritis UK Centre for Genetics and Genomics, Centre for Musculoskeletal Research, University of Manchester, Manchester, UK; Department of Clinical Sciences, Lund University, Helsingborg, Sweden; Precision Healthcare University Research Institute, Queen Mary University of London, London, UK; Division of pulmonary and critical care medicine, Mass General Brigham, Boston, MA; Section on Pulmonary, Allergy, Critical Care, and Sleep Medicine, Department of Veterans’ Affairs, West Roxbury, MA

**Keywords:** obesity, proteomic score, BMI, multi-ancestry, generalizability, cardiometabolic

## Abstract

Plasma proteomic scores for body mass index have been derived primarily in European populations, and it is unclear whether these scores are generalizable and useful in other ancestries. Here, we show that plasma proteomic BMI scores developed in participants of European, East Asian, South Asian and African ancestries from the UK Biobank (UKBB; N=50,621) and the South African Middle-Aged Soweto Cohort (MASC; N=948) are portable across ancestries, explain up to 48.7% of trait variance and significantly predict incident obesity. We identified eight protein biomarkers common across all scores, six of which showed causal associations with BMI in Mendelian randomization analyses and were associated with BMI across the Olink (UKBB, MASC) and SomaScan platforms (Qatar Biobank; N= 2410). Discrepancy analysis between the high predicted proteomic BMI and low measured BMI revealed metabolically unhealthy normal weight (MUNW) individuals, with higher visceral fat, elevated triglycerides, and low insulin sensitivity. Thus, plasma proteomic BMI scores may enhance the precision of cardiometabolic risk stratification across diverse populations.

---

Obesity is a modifiable risk factor for cardiometabolic diseases, and it is projected to affect 1.1 billion adults globally by 2030, representing a 110% increase from 2010 ^1^. The associated non-communicable diseases (NCDs), including cardiovascular diseases and type 2 diabetes, pose a public health crisis as they are associated with premature disability, death and increased health care spending ^2–4^. Body mass index (BMI) has traditionally served as a metric for obesity screening and classification at the population level ^5^. While it shows high correlations with body fat percentage and visceral fat derived from dual X-ray absorptiometry (DXA) and computed tomography (CT), its associations with health outcomes are heterogeneous. Individuals with the same BMI can have varied cardiometabolic risk profiles, limiting its utility at an individual level ^6–10^.

Multiple approaches are recommended to enhance the utility of BMI in NCD risk stratification, including supplementing it with anthropometric measures such as waist circumference and multi-omic profiles^11,12^. However, waist circumference thresholds are more Eurocentric and have limited diagnostic accuracy in diverse populations^13,14^. Above all, anthropometric measures may not capture the underlying metabolic risk and misclassify people as normal weight who may be ‘metabolically obese’ and at a higher risk of CMD and mortality^15,16^. There is now a growing interest in using BMI omic profiles derived from proteomics, genetics and metabolomics in cardiometabolic risk prediction ^12,17,18^. Omic profiles offer additional biological insights, capturing tissue-specific and age-dependent metabolic processes that BMI alone does not reflect ^19,20^. Discordance between measured BMI and omics-predicted BMI, particularly among individuals classified as normal weight by BMI but with obesity by their omics profile, has been associated with elevated risks of mortality and cardiometabolic disease in European populations ^15,21^. These molecular signatures may help explain the heterogeneity in metabolic health observed among individuals with similar BMI ^22–25^. In addition, plasma/serum-derived proteomic profiles have identified several proteins involved in metabolic pathways, oxidative stress and inflammatory responses, that are altered in obesity and related disease conditions ^26–28^. However, the generalizability of these associations to people of other ancestries remains unclear.

Because proteomic and profiles are not constrained by linkage linkage disequilibrium (LD) or allele frequency, they might be more generalizable than polygenic risk scores ^12,29,30^. Proteomic scores have been observed to account for up to 70% of BMI trait variance, which is 4.5 times higher than has been recently reported for PRSs using the largest BMI GWAS discovery of 5 million participants of European ancestry (PRS R^2^ = 17.6%) ^31^. However, many of these proteomic studies have been conducted in populations of European ancestry, and there is limited evidence of the cross-ancestry generalizability of the derived scores ^32,33^. Establishing cross-ancestry generalizability of proteomic scores holds promise for improving risk stratification and providing insights into the pathways associated with BMI and obesity-related cardiometabolic disease (CMD) outcomes worldwide ^34,35^. In this analysis, we leveraged longitudinal proteomic and phenotypic data available from the South African Middle-Aged Soweto cohort (MASC) and multiple ancestry groups in the UK Biobank (UKBB) to evaluate the cross-ancestry predictivity of proteomic scores for BMI and incident obesity over a nine-year period. In addition, we compared the predictive values of both proteomic scores and measured BMI for CMD outcomes. Finally, we evaluated the cross-platform (Olink to SomaScan) generalizability of a proteomic BMI score based on a parsimonious set of proteins and assessed its utility for identifying the metabolically unhealthy normal weight (MUNW) phenotype in the Middle Eastern Qatar Biobank (QBB) participants.

## Results

### Computation of plasma proteomic scores

To develop BMI proteomic signatures, we computed weighted plasma proteomic scores for BMI in 51,569 individuals from multiple ancestries (South Africans of African ancestry in MASC – contAFRPPS; East Asians - EASPPS, South Asians - SASPPS, Africans - diaAFRPPS and Europeans – EURPPS in the UKBB) using adaptive LASSO with 10 fold cross validation in the training data set (70%) and leave out analysis in the test data set (30%) as indicated in the overview of the study design (**Figure 1, Methods**). Ancestry was determined using genetic principal components in the UKBB. Participants were middle-aged (MASC: 53 ± 5.9 years; UK Biobank 56.9 ± 8.2 years) with similar mean BMI (29.7 ± 5.4 kg/m^2^(MASC); 29.7 ± 7.5 kg/m^2^ (UKBB)) (**Supplementary Table 1**). The contAFRPPS comprised 91, EURPPS 149, diaAFRPPS 66, EASPPS 19 and SASPPS 74 proteomic biomarkers (**Figure 3, Supplementary Table 2**). All proteomic scores were significantly associated (p<0.05) with BMI (**Figure 2A, Supplementary Table 3**) and explained the BMI trait variance ranging from 55.3% (EASPPS) to 67.6% (contAFRPPS) in their respective ancestry groups (**Figure 2B, Supplementary Table 3**).

**Figure 1:**
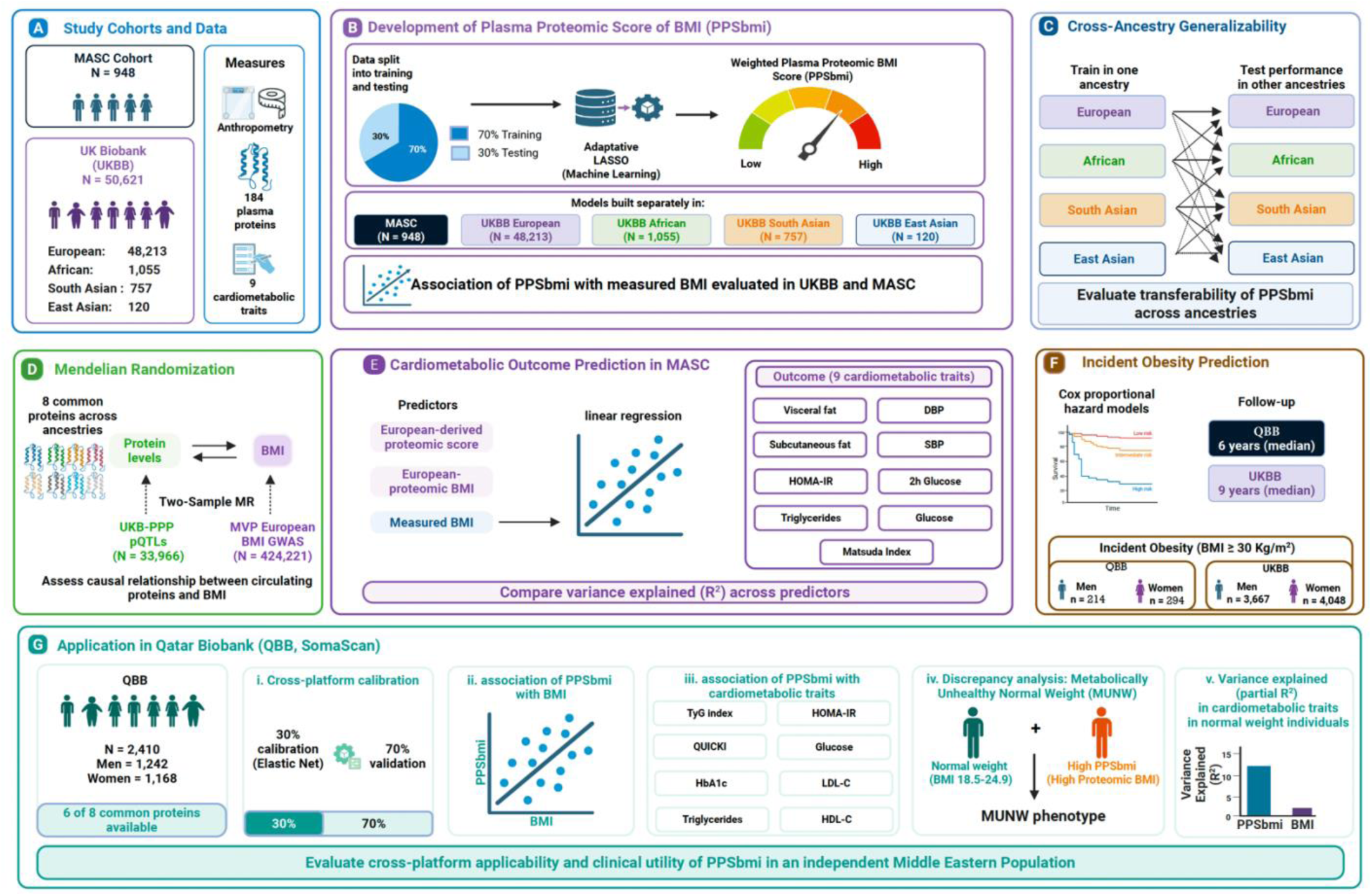
Study design overview. **A:** Description of the study cohorts. **B:** Development of the weighted proteomic score for BMI in the MASC cohort and multiple genetic ancestry populations of the UKBB cohort (East Asians, South Asians, Africans, and Europeans) using a machine-learning approach (adaptive lasso). **C**: Evaluation of the cross-ancestry generalizability of the BMI proteomic scores. **D**: Two-sample Mendelian Randomization (MR) analysis of 8 proteins that were common across all ancestries. **E:** Linear regression modelling to assess association of EUR proteomic scores, EUR proteomic BMI and measured BMI with the selected CMD markers in MASC. **F:** Prediction of incident obesity using Cox proportional hazards models in MASC and European-ancestry UKBB participants who had at least two BMI measurements and proteomic data. **G.** Cross platform application of BMI-eight-proteomic score in the Qatar Biobank (QBB).

**Figure 2:**
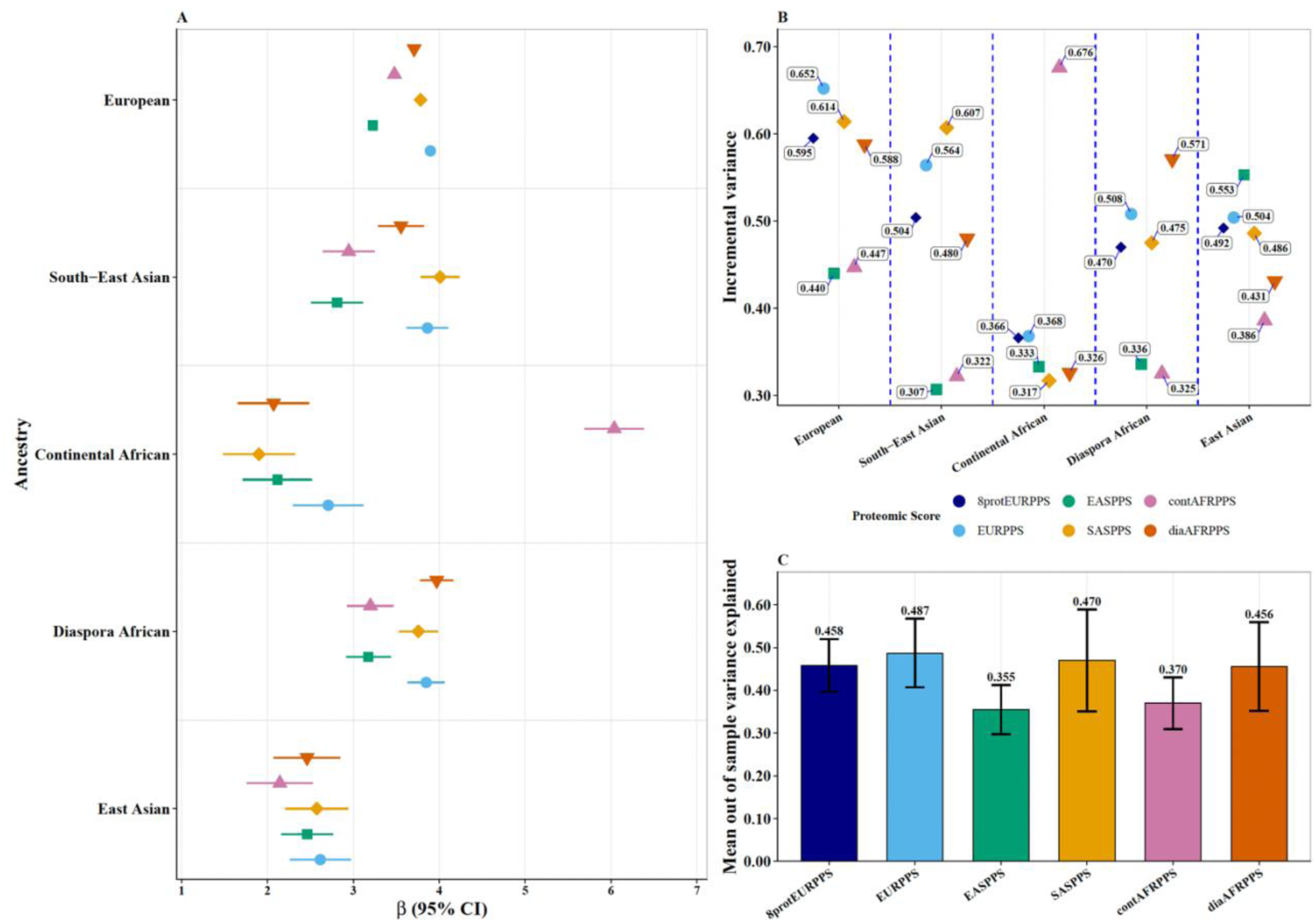
Cross-ancestry associations of proteomic scores derived in the UK Biobank (UKBB) and the Middle-Aged Soweto Cohort (MASC) with BMI. **A:** Forest plot showing the effect sizes of the BMI proteomic scores across ancestry groups in the UKBB and MASC. **B:** Incremental trait variance explained by the BMI proteomic scores across the ancestries in the UKBB and MASC. **C:** Mean out of sample trait variance in BMI explained by proteomic scores. Out-of-sample refers to variance explained in other ancestries excluding the ancestry in which the score was derived from. Error bars represent 95% confidence intervals.

**Figure 3A:**
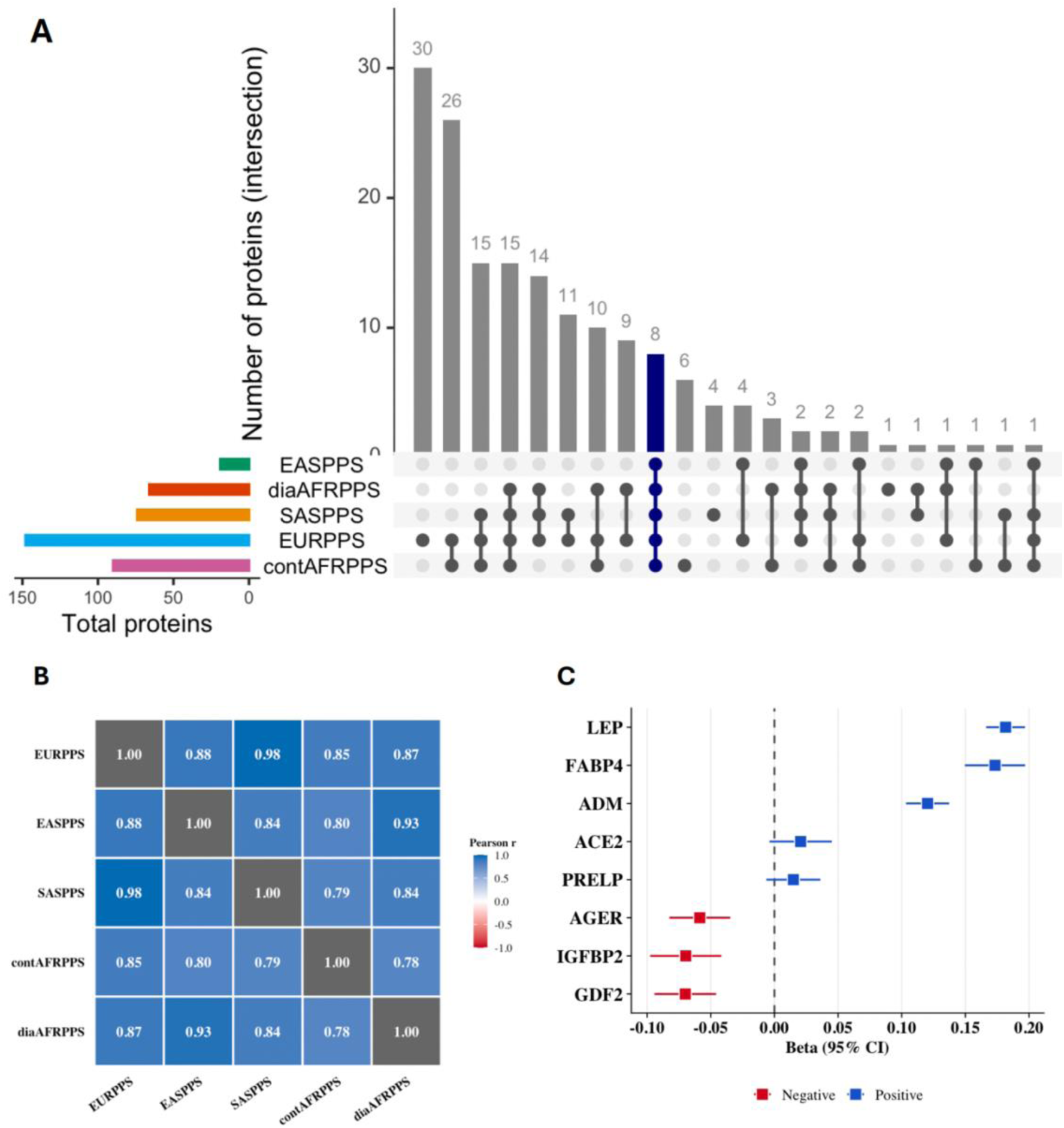
Shared proteomic biomarkers. Venn diagram showing overlap of proteomic biomarkers across the five proteomic scores derived in the UK Biobank (UKBB) and the Middle-Aged Soweto Cohort (MASC) with BMI. Proteomic scores for BMI were derived in multiple self-reported ancestries (MASC - contAFRPPS, East Asian ancestry - EASPPS, South Asian ancestry - SASPPS, African ancestry in the UKBB - diaAFRPPS and European ancestry - EURPPS) using a machine learning method (adaptive lasso) The contAFRPPS comprises 85 proteomic biomarkers, UKBB-derived scores comprise, 152 (EURPPS), 66 (diaAFRPPS), 18 (EASPPS) and 70 (SASPPS) biomarkers. Eight biomarkers (ADM, AMBP, FABP4, GDF2, IGFBP2, LEP, MERTK and AGER) are shared across all five scores. **B.** Pairwise correlations of the adaptive-lasso effect sizes for the eight shared biomarkers in proteomic scores derived in multiple ancestries. Proteomic scores were derived in the Middle-Aged Soweto Cohort (MASC – contAFRPPS) and the UKBB ancestry groups: East Asian ancestry - EASPPS, South Asian ancestry - SASPPS, African ancestry - diaAFRPPS and European ancestry – EURPPS. Correlations were obtained using the pheatmap_1.0.13 package of R (version 4.4.2) and strength is shown as a gradient from low (light blue) to high (dark blue). **C.** Mendelian randomization (MR) analysis estimating the causal effect of BMI on eight proteomic biomarkers identified as shared across diverse-ancestry proteomic scores derived in the UK Biobank (UKBB) and MASC cohorts. MR analysis was conducted using a random-effects inverse-variance weighted (IVW) approach implemented in TwoSampleMR v0.5.6. Genetic instruments for BMI were obtained from the Million Veteran Program (MVP) European ancestry BMI GWAS (N=424,221) and protein outcomes from the UK Pharma Proteomics Project (UKB-PPP) (Nmax=33,966).

### Shared proteomic biomarkers across the BMI proteomic scores

We then sought to determine the shared proteomic biomarkers across the five ancestry-specific proteomic scores and genetic evidence of their causal association with BMI. We identified eight proteomic biomarkers (ACE2, ADM, FABP4, GDF2, IGFBP2, LEP, PRELP, and AGER) that were common to all proteomic scores (**Figure 3A**). Six of these proteomic biomarkers had concordant LASSO model coefficient directions across all scores except for ACE2 and PRELP. ACE2 was positively associated with BMI in all ancestries but negatively associated in South Africans of African ancestry. PRELP was negatively associated with BMI in South Africans of African ancestry, UKKB Africans, and UKBB East Asians, whereas it was positively associated with BMI in other ancestry ancestries (**Supplementary Table 2**).

We then evaluated the correlations of the eight common proteomic biomarkers using the ancestry specific LASSO model coefficients. The coefficients of the eight proteomic biomarkers strongly correlated across ancestries there were derived from, with correlation coefficient (R) ranging from 0.78 to 0.98 (**Figure 3B**).

We proceeded to determine the genetic evidence of causal association of the eight shared proteomic biomarkers and BMI. We examined each causal direction (protein → BMI and BMI → protein) separately, utilizing the Million Veterans Program (MVP) GWAS of BMI (N = 424,221) in individuals of European ancestry and protein outcomes from UK Pharma Proteomics Project (UKB-PPP) (N = 33,966) **(Methods)** ^36,37^. We found that BMI had a causal effect on the expression of six of the eight proteins. A higher BMI was associated with lower expression of three proteins (IGFBP2, AGER and GDF2) and higher expression of LEP, FABP4 and ADM. (**Figure 3C, Supplementary Tables 5-16**). Subsequently, using the cis-pQTLs (when available) identified in the UKB-PPP ^38^ as instruments for those same protein biomarkers we performed reverse MR and found evidence of reverse causality only for AGER. Thus, the expression of the six proteins is causally influenced by BMI and AGER expression increases BMI **(Supplementary Tables 7 and 8)**.

### Cross-ancestry generalizability of the BMI proteomic scores

We then sought to evaluate the cross-ancestry generalizability of each score by applying the score to other ancestries, except the one from which it was derived and then obtaining the mean BMI trait variance it explained across ancestries (**Figure 2C**). We found that all the scores were portable to other ancestries, with a mean out-of-sample trait variance (R^2^) ranging from 35.6% (EASPPS) to 48.7% (EURPPS). (**Figure 2, Supplementary Table 4**). Notably, the BMI proteomic score comprising eight shared proteomic biomarkers explained almost similar mean BMI trait variance (R^2^ = 45.8%) across the ancestries as the full EURPPS (**Figure 2C**, **Supplementary Table 4**).

### Associations of proteomic scores with cardio-metabolic traits

After observing the cross-population generalizability of the BMI proteomic scores, we sought to determine the association between these scores with cardiometabolic outcomes in MASC participants independent of BMI or waist circumference. Cardiometabolic traits were scaled for easier comparative analysis in the forest plots (Methods). We used the European ancestry proteomic score (EURPPS) and 8-protein score (8protEURPPS) as predictors in our models. Age, sex were used as covariates together with BBMI and waist circumference in stratified analysis **(Figure 4, Methods)**. Notably, the 8protEURPPS and EURPPS showed similar magnitudes of association and were independently correlated with VAT, SAT, triglycerides and insulin sensitivity (Matsuda index), thereby showing the utility of these BMI proteomic scores in capturing additional variance in these traits that are independent of BMI and waist circumference (**Supplementary Table 17**).

**Figure 4:**
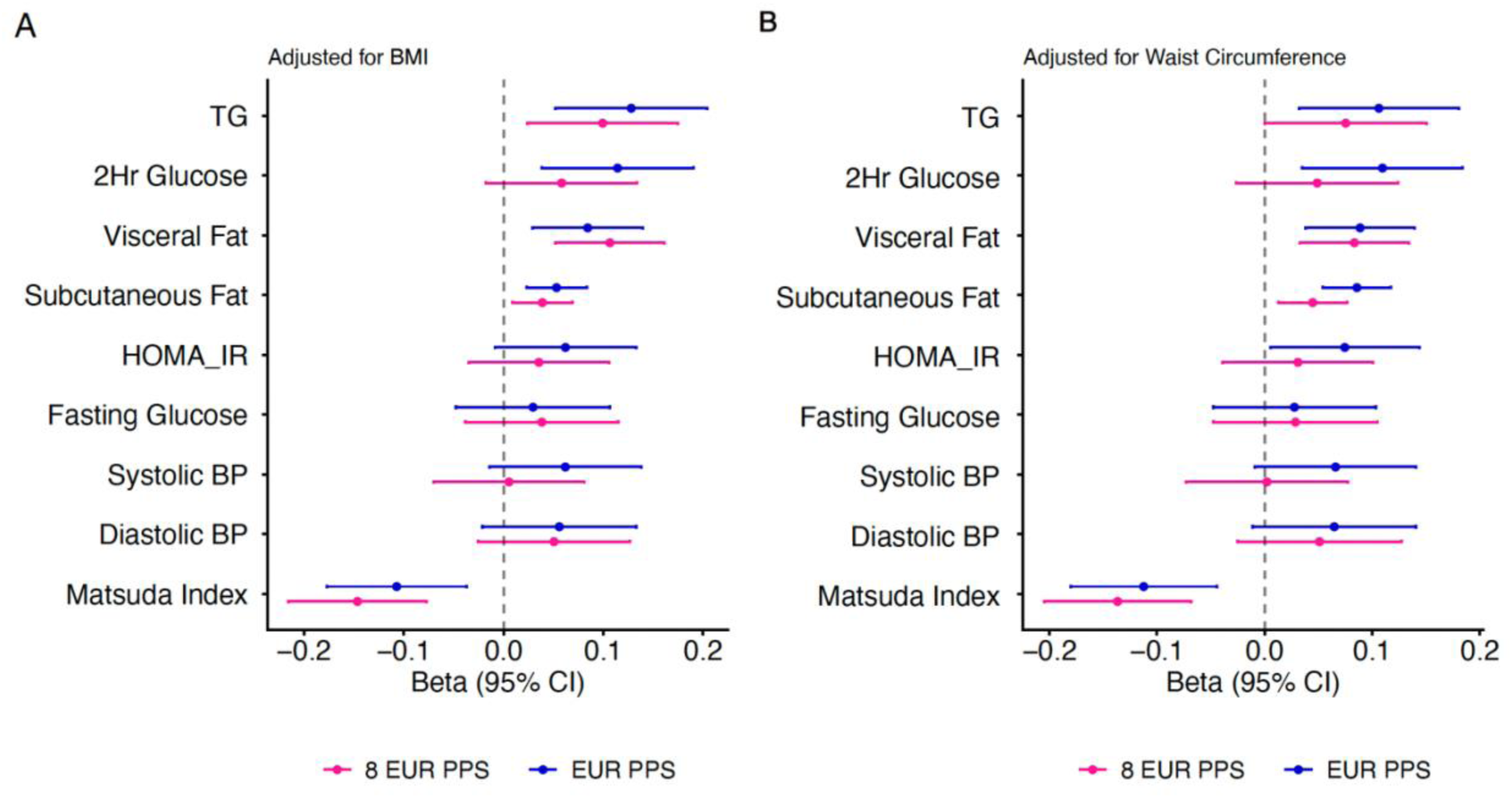
Association of proteomic scores and proteomic-inferred BMI with cardiometabolic outcomes among South Africans of African ancestry in the MASC cohort. **A:** Association of two proteomic scores adjusted for age, sex and BMI **B:** Associations of two proteomic predicted BMI (EURPPS and EUR8PPS) with selected cardiometabolic outcomes adjusted for waist circumference. Multiple testing correction was done using the Benjamini–Hochberg false discovery rate (FDR) procedure.

### Cross-platform recalibration in QBB

Finally, we assessed whether our results could be replicated in an independent population profiled using a different proteomic platform. Owing to the differences in proteomic platforms between MASC and UK Biobank versus QBB, we evaluated the cross-platform generalizability of the parsimonious 8protEURPPS. Of the eight overlapping proteins, six (LEP, GDF2, AGER, IGFBP2, ACE2 and ADM) were available in QBB and were used to derive the proteomic BMI score following recalibration in QBB using Elasticnet (Methods). The model coefficients retained the same direction of effect as those observed in the 8protEURPPS (**Supplementary Table 18**).

Similar to the results observed in other ancestries, the recalibrated six-protein plasma proteomic score (6protPPS) was correlated with the measured BMI and explained 46% of the trait variance (Pearson’s *r* = 0.67, *P* < 0.001; **Figure 5A**). We investigated associations between the 6protPPS predicted BMI (6protpBMI) with measures of insulin resistance: HOMA-IR, triglyceride glucose Index (TyG Index); a measure of insulin sensitivity: quantitative insulin sensitivity check index (QUICKI); glucose, glyacaemic control (HbA1c); triglycerides, total cholesterol, HDL Cholesterol and LDL Cholesterol.

**Figure 5.**
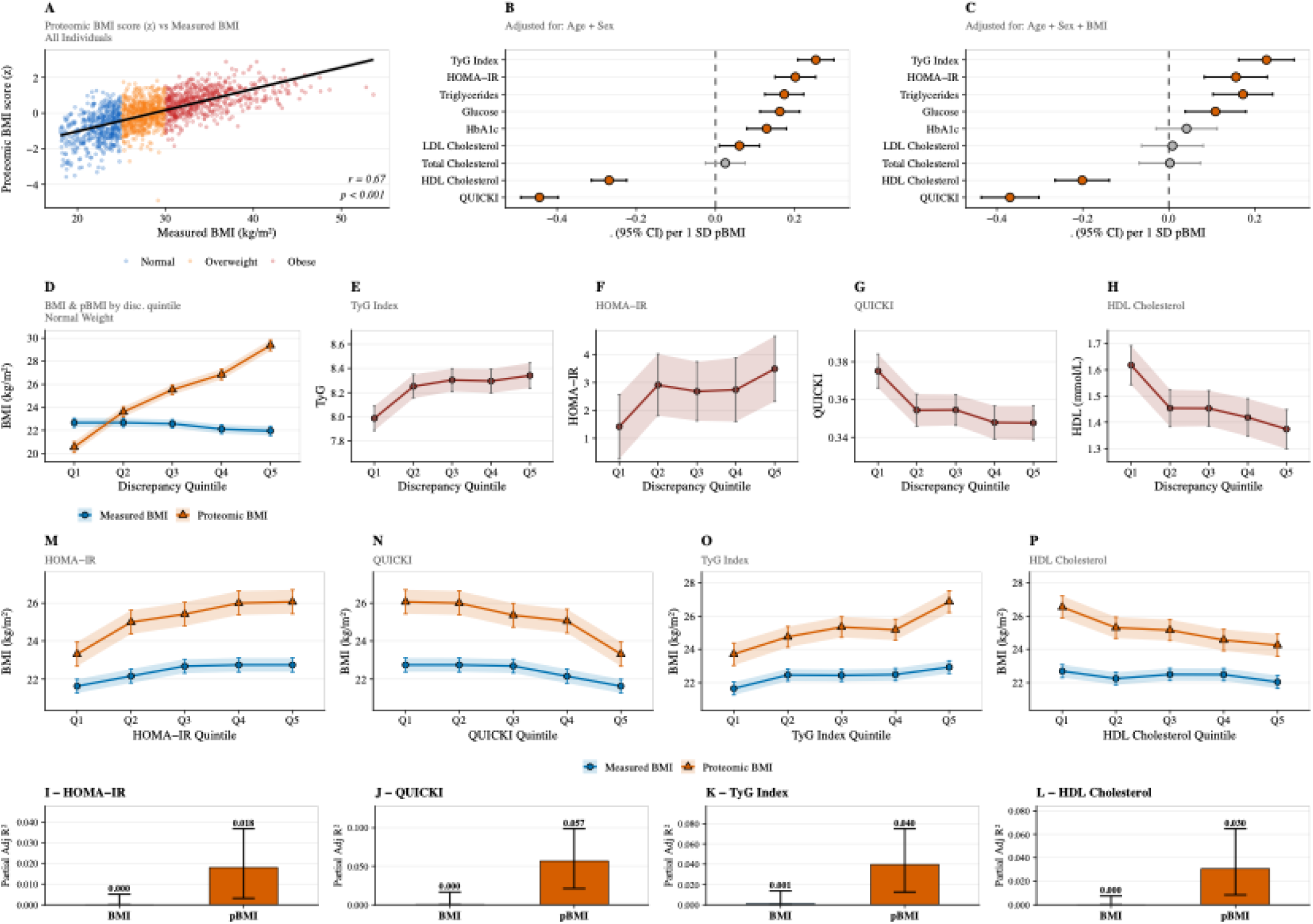
Proteomic BMI (pBMI) in Qatar Biobank Cohort. **A.** Correlation between the proteomic BMI score (z-scored) and measured BMI in QBB validation set individuals (n = 1,685). **B–C.** Forest plots showing standardised regression coefficients (β ± 95% CI) for the association between the proteomic BMI (pBMI, 1 SD increment) and cardiometabolic traits in QBB validation set, adjusted for age and sex only **(B)** or additionally adjusted for measured BMI **(C)**. **D.** Estimated marginal means (± 95% CI) of measured BMI and proteomic BMI (pBMI) across discrepancy quintiles (pBMI − BMI) in Normal Weight individuals (n = 461), adjusted for age and sex. **E–H.** Estimated marginal means (± 95% CI) of TyG index **(E)**, HOMA-IR **(F)**, QUICKI **(G)**, and HDL cholesterol **(H)** across discrepancy quintiles in Normal Weight individuals, adjusted for age and sex. **M–P.** Estimated marginal means (± 95% CI) of measured BMI and proteomic BMI across quintiles of HOMA-IR **(M)**, QUICKI **(N)**, TyG index **(O)**, and HDL cholesterol **(P)** in Normal Weight individuals, adjusted for age and sex. Quintiles are defined within the Normal Weight stratum. **I–L**. Partial adjusted R² of measured BMI and proteomic BMI for HOMA-IR **(I)**, QUICKI **(J)**, TyG index **(K)**, and HDL cholesterol **(L)** in Normal Weight individuals. Partial R² quantifies the unique variance explained by each predictor after adjusting for the other, from a model including age, sex, BMI, and pBMI. Error bars represent 95% percentile confidence intervals derived from 1,000 bootstrap resamples of the Normal Weight validation set. All R² values are adjusted R².

Independent of measured BMI, higher proteomic BMI was associated with greater insulin resistance and a more adverse cardiometabolic profile (**Figure 5B**, **5C; Supplementary Table 19**). Cardiometabolic traits were scaled for comparison on the same scale in the forest plots. Specifically, each 1-SD increase in proteomic BMI was associated with higher TyG index (β = 0.23, 95% CI: 0.16 - 0.29; *P* = 8.65 × 10⁻¹²), HOMA-IR (β = 0.16, 95% CI: 0.08 - 0.23; *P* = 3.03 × 10⁻⁵), and lower QUICKI (β = −0.37, 95% CI: −0.44 to −0.30; *P* = 3.37 × 10⁻²⁶). Higher proteomic BMI was also associated with elevated triglyceride levels (β = 0.17, 95% CI: 0.10 - 0.24; *P* = 1.22 × 10⁻⁶), lower HDL cholesterol levels (β = −0.20, 95% CI: −0.26 to −0.14; *P* = 5.02 × 10⁻¹⁰) and higher glucose levels (β = 0.11, 95% CI: 0.04 - 0.18; *P* = 0.003). These associations were independent of BMI (**Figure 5C, Supplementary Table 22**).

### Discrepancy between measured BMI and proteomic BMI in normal-weight individuals in QBB

Here, we evaluated the discrepancy between the 6protpBMI and measured BMI to identify individuals who are classified as normal weight by BMI but exhibit an overweight or obese proteomic profile. Among normal-weight individuals (BMI < 25 kg/m², n = 461), the measured BMI remained relatively stable across quintiles of the scaled differences in BMI discrepancy (6protpBMI - BMI), whereas 6protpBMI reach almost obese levels of 29.4 kg/m² in the highest quintile (Q5) **(Figure 6D)**. This discrepancy was also associated with unfavorable metabolic profiles. Compared to individuals in Q1, those in Q5 exhibited a significantly higher TyG index (Q5: 8.34 [8.24, 8.45] vs Q1: 7.987 [7.88, 8.09]; p = 1.17 × 10⁻⁵), higher HOMA-IR (Q5: 3.49 [2.32, 4.65] vs Q1: 1.41 [0.26, 2.56]; p = 0.022), lower QUICKI (Q5: 0.35 [0.34, 0.36] vs Q1: 0.38 [0.37, 0.38]; p = 9.99 × 10⁻⁵) and lower HDL cholesterol levels (Q5: 1.37 [1.30, 1.45] vs Q1: 1.62 [1.54, 1.69] mmol/L; p = 3.31 × 10⁻⁵) (**Figure 5E–H; Supplementary Table 20-21**).

**Figure 6.**
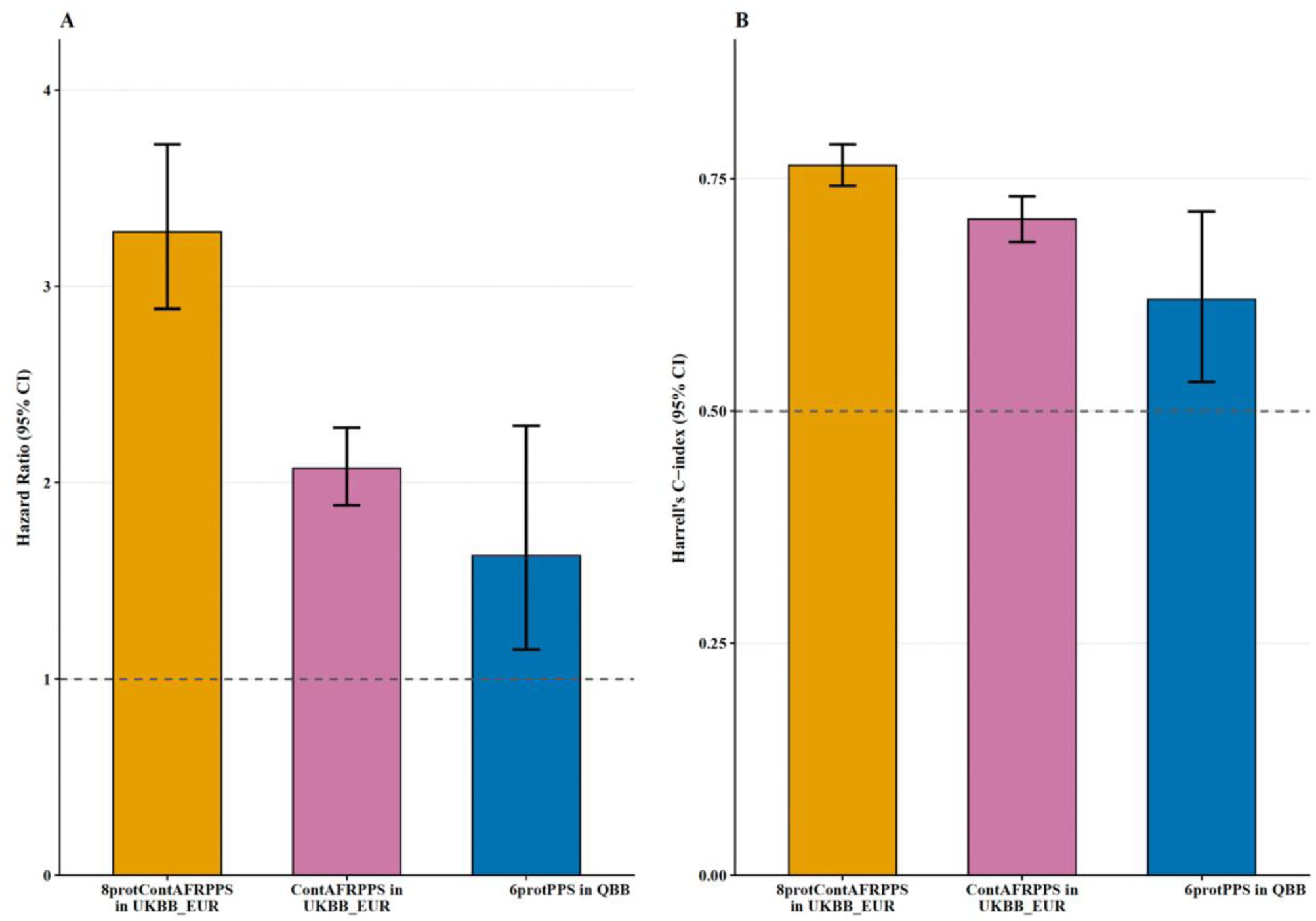
Proteomic prediction of incident obesity in the UK Biobank and QBB. The predictivity of incident obesity by the full contAFRPPS and 8 protContAFRPPS in the UKBB over a 9-year period; then 6protPPS in the QBB over a 6-year period using Cox proportional hazards models. The results are presented as **A.** Hazard Ratios, 95% CI and **B.** C-statistics and 95%CI

Furthermore, we evaluated whether the 6protpBMI more sensitively captures metabolic deterioration among normal-weight individuals across HOMA-IR, TyG index, QUICKI and HDL cholesterol quintiles. 6protpBMI exhibited consistently steeper gradients than measured BMI, indicating greater sensitivity to worsening metabolic profiles (**Figure 5I–L**). For HOMA-IR, the positive linear trend was more than two-fold stronger for 6protpBMI than measured BMI (β = 0.65, P = 3.16 × 10⁻¹⁰ vs. β = 0.28, P = 2.17 × 10⁻⁶). Similarly, 6protpBMI increased more markedly across TyG index quintiles than measured BMI (β = 0.68 vs. 0.26; P = 7.32 × 10⁻⁹ vs. 1.49 × 10⁻⁴). Conversely, 6protpBMI declined more sharply across increasing QUICKI quintiles (β = −0.648 vs. −0.284; P = 5.16 × 10 ⁻¹⁰ vs. 2.00 × 10⁻⁶). A similar pattern was observed for HDL cholesterol, where 6protpBMI demonstrated a significant inverse trend (β = −0.53, P = 3.95 × 10⁻⁶), whereas measured BMI showed only a weak, non-significant association (β = −0.10, P = 0.122). Collectively, these findings indicate that 6protpBMI is more responsive to progressive metabolic deterioration than conventional BMI, suggesting that it more sensitively captures the underlying metabolic dysfunction in individuals classified as normal weight.

### Variance of metabolic traits explained by proteomic BMI in normal-weight individuals

Among normal-weight individuals, 6protpBMI consistently explained a greater proportion of trait variance in markers of metabolic health than measured BMI. Using partial variance in which both measures were included in the same model, the unique contribution of BMI was negligible across all traits, with partial R² values close to zero. In contrast, 6protpBMI retained independent explanatory value for HOMA-IR (partial R² = 0.02, 95% CI: 0.003 −0.04), QUICKI (partial R² = 0.06, 95% CI: 0.02 - 0.10), TyG index (partial R² = 0.04, 95% CI: 0.01 - 0.08), and HDL cholesterol (partial R² = 0.03, 95% CI: 0.01 - 0.06) (**Figure 5M-P, Supplementary Table 22**). Overall, these findings indicate that proteomic predicted BMI captures metabolic variation beyond that reflected by conventional anthropometric BMI, even among individuals with normal weight.

### Longitudinal analysis

We further assessed the utility of proteomic scores in predicting incident obesity. We fitted Cox proportional hazards models with incident obesity (BMI ≥ 30kg/m^2^) as the outcome, excluding participants with obesity at baseline **(Figure 1F, Methods)**. Due to sample size constraints for other UKBB ancestries (fewer than 100 participants) this analysis focused only on participants of European ancestry (UKBB) and QBB (**Methods, Supplementary Table 23-24**). A cross-population analysis was conducted to evaluate the generalizability of longitudinal prediction of the scores by using contAFRPPS scores to predict incident obesity in Europeans in the UKBB. We observed similar predictive performance of the for the standard deviation increase in the full contAFRPPS (Harrel’s C-static 0.71, 95%CI(0.68-0.73)) and the 8 protcontAFRPPS (Harrel’s C-static 0.76, 95%CI(0.74-0.79)) of incident obesity in Europeans in the UKBB (**Figure 6, Supplementary Table 20**). The 6protPPS significant was predicted incident obesity in the QBB (Harrel’s C-static 0.62, 95%CI(0.53-0.72))

## Discussion

Our findings demonstrate that proteomic BMI scores explain 55% to 67% of BMI trait variance in their respective discovery ancestries and generalize well across populations, showing concordant effect sizes and explaining up to 48.7% of BMI variance, substantially more than is typically reported for PRS ^31,39,40^. Moreover, we observed that eight proteins (ACE2, ADM, FABP4, GDF2, IGFBP2, LEP, AGER and PRELP) were shared across all scores, with six of them having evidence of genetic causal associations through Mendelian randomization. In addition, the proteomic score derived from these eight common proteins explained 45.8% of BMI variance across ancestries and had similar predictive performance to the 149 proteomic marker score (EURPPS). Further, the proteomic score explained greater variance in VAT, estimated insulin sensitivity (Matsuda Index) and TG than measured BMI in South Africans of African ancestry. This eight-protein score was portable across platforms of Olink and Somascan and was useful in the detection of metabolically unhealthy normal weight (MUNW) people. Together, these results underscore the generalizability and predictive robustness of these scores across diverse populations, highlighting their potential utility for understanding obesity risk and CMD across diverse populations.

The ancestry-derived proteomic scores of BMI explained the BMI trait variance from 55% - 67% of the BMI trait variance in their discovery ancestries. These predictions were similar to the 70% BMI trait variance reported previously in European populations^12^. This illustrates the benefits of proteomic scores compared to genetic risk scores, in that with just a discovery dataset of 120 in East Asians, the BMI trait variance could be explained more than using a very large discovery dataset in genetics at the scale of 5 million to explain 17% of the BMI trait variance in Europeans and 2% in continental Africans^41^. Although mean trait variance explained across populations, was lower than the ancestry specific scores (35.6% for EASPPS to 48.7% for EURPPS), it remained substantially higher than typically reported for PRS predictions. Notably, the parsimonious 8protEURPPS of the shared proteins across ancestry scores had a mean trait variance of 45.8% thus further showing the potential utility of scaling and applying very few proteomic biomarkers across ancestries while accounting for BMI trait variance better than polygenic risk scores. This is largely because proteomic biomarkers are universal and not limited by differences in LD and allele frequencies, as in the genetic variants in PRS ^42^. Moreover, the strong correlations (R = 0.75 to 0.98) between the coefficients of the eight proteins shared across all five ancestries and causally linked to BMI further supports the existence of conserved biological pathways underlying BMI-associated metabolic regulation, despite ancestry-specific differences ^17,33,43^.

In our analysis, the proteomic score was associated with key measures of adiposity and cardiometabolic risk, including VAT, SAT, TG, insulin sensitivity, blood pressure and glycaemic traits mirroring associations observed with measured BMI. Additionally, proteomic BMI explained additional trait variance in VAT, insulin sensitivity and TG beyond that explained by measured BMI or waits circumference in South Africans of African Ancestry in MASC and Qatari people in the QBB. This suggests that proteins within these score may capture a stronger signal of metabolic dysfunction beyond anthropometric assessments. Consistent with our results, prior studies have reported associations between proteomic predicted BMI and cardiometabolic traits. However, these studies were conducted in predominately European populations and did not assess important traits such as insulin sensitivity and VAT ^12,17^. Our observations expand on previous findings, suggesting that proteomic BMI captures biological processes related to metabolic dysfunction that extend beyond adiposity alone, especially those linked to insulin sensitivity.

The proteomic scores were found to be useful for predicting incident obesity and for risk stratification. In the longitudinal analysis, the Harrel’s C-index ranged from 0.59 to 0.72, with hazard ratios 1.88 - 4.32. Similarly, increases in Harrel’s C-indices and hazard ratios have been reported for other traits in European populations ^32,44^. Importantly, we show improved predictive power and risk stratification of proteomic scores is maintained in QBB and UKBB populations, in contrast to the limited portability reported for PRS. The similar predictions of the 8contAFRPPS and the full contAFRPPS with 91 proteomic biomarkers show the potential to translate and test the parsimonious score in future studies at scale in diverse populations.

A major challenge for proteomics-based risk scores is limited transferability across analytical platforms, as differences in protein coverage and quantification often hinder external validation ^45^. Despite being derived from only six proteins shared across platforms, the 6protpBMI score generalized well to the Qatar Biobank, an independent SomaScan-based cohort representing a Middle Eastern population. The ability of this parsimonious protein panel to retain strong associations with metabolic traits across both the O-link and Somascan platforms and different populations suggests that these proteins capture the fundamental biological processes underlying adiposity and metabolic dysfunction rather than platform-specific measurement artefacts. Importantly, proteomic BMI remained associated with adverse metabolic traits, particularly insulin resistance, independent of measured BMI. These findings extend previous observations from Asian populations and support the broader generalizability of obesity-related proteomic signatures across diverse ancestral and environmental backgrounds ^17^.

Beyond replication, our results provide biological insight into the discrepancy between proteomic and measured BMI. We showed that discordance between the two measures is not merely measurement noise but reflects meaningful metabolic heterogeneity. Previous studies have reported similar observations using metabolomics-derived BMI in Europeans, showing that the discordance between omics-based BMI and measured BMI captures metabolic heterogeneity that is not reflected by anthropometric measures alone ^15,21^. However, circulating proteins are generally considered more stable biological markers and have been shown to be more resistant to short-term lifestyle-induced changes than metabolites ^12^. Consequently, proteomic BMI may provide a more robust measure of an individual’s underlying metabolic state and long-term disease risk. In our study, individuals with a higher proteomic BMI than expected for their measured BMI exhibited progressively worsening insulin resistance and adverse cardiometabolic profiles despite similar anthropometric status, as observed in the discrepancy analysis, whereas those with lower proteomic BMI appeared metabolically healthier. Importantly, this information was captured using a parsimonious six-protein panel, highlighting the potential utility of proteomic BMI as a scalable biomarker for identifying individuals whose metabolic risk may be underestimated by conventional anthropometric measurements. These findings are consistent with the MUNW phenotype ^8^ and suggest that proteomic profiling can identify metabolically vulnerable individuals who would be overlooked by conventional anthropometric screening.

Collectively, our findings support the view that complementing anthropometric data with proteomic informed obesity phenotyping is pivotal for cardiometabolic risk. Although BMI remains widely used because of its simplicity and low cost, it does not adequately capture inter-individual differences in metabolic health. By integrating molecular information, proteomic BMI-based discordance measures may identify biologically vulnerable individuals who would otherwise be misclassified using conventional BMI criteria alone. The ability of a six-protein panel to detect metabolic heterogeneity and identify normal-weight individuals with adverse metabolic profiles highlights the potential for scalable blood-based approaches such as dry blood spot to improve cardiometabolic risk stratification, particularly in resource-limited settings where advanced imaging modalities for assessing body composition and ectopic adiposity are not readily available.

This analysis has several strengths, including the use of multi-ancestry cohorts with longitudinal follow-up in two ancestries, robust cross- and multi-population validation, comprehensive proteomic characterization and sex-stratified analysis. However, there are some limitations to consider, including the limited number of proteomic biomarkers evaluated (only the CVDII and CVDII O-link panels were used) and the absence of longitudinal data for ancestry populations other than continental Africans and Europeans.

## Conclusion

This study demonstrates the portability of plasma derived proteomic scores for BMI across diverse ancestries, despite differences in genetic architecture. We also identified shared proteomic biomarkers in the ancestry specific scores that are genetically causally associated with BMI. These findings reflect the potential to scale and enhance the use of proteomic BMI as a complementary measure to anthropometric BMI, using only a few proteomic markers. In adults, proteomic BMI appears more sensitive than BMI to visceral adiposity and insulin sensitivity, suggesting added cardiometabolic risk stratification potential. However, it remains to be determined whether similar patterns hold in children, adolescents and older adults.

## Methods

### Study participants

Our study participants were from the Middle-age Soweto Cohort (MASC) in South Africa (N = 984), the UK Biobank (UKBB) in UK and the Qatar Biobank (QBB) in Qatar. The MASC is a nested study of the Africa Wits-INDEPTH partnership of Genetic Research (AWI-Gen) cohort and includes black South African men and women aged between 29 to 82 years at baseline. AWI-Gen is a longitudinal cohort from four African countries^13,46^ in three geographical regions. The UKBB is, a large-scale prospective cohort of over 500,000 individuals aged 40 years and above at baseline from the United Kingdom^47^. Of the 500,000 individuals recruited in the UKBB, 50,621 have proteomic data and these individuals were stratified into European, East Asian, South Asian and African ancestries using genetic principal components. QBB is a population-based initiative under Qatar Precision Health Institute (QPHI) to promote biomedical research in Qatar and worldwide ^48^. So far, the QBB has recruited 2,410 nondiabetic men and women aged 18 to 80 years. The recruitment was restricted to Qatari citizens and long-term residents (≥15 years living in Qatar) who were followed up every 5 years to get update on their health status. All study participants have proteomic data collected at baseline and available for analysis.

### Ethical clearance

Ethical clearance to conduct this study was obtained from the University of Witwatersrand Human Research Ethics Committee (Protocol Number: M210166) and Mass General Brigham IRB (Protocol Number: 2021P0003536). Ethical clearance for the UKBB is obtained on a 5-year cycle and the current approval with reference 21/NW/0157 was granted by the Northwest - Haydock Research Ethics Committee. Ethical approval was obtained by the Institutional Review Boards of Qatar Biobank (QBB; Approval No. E-2019-QBB-RES-ACC-0179-0104) and Hamad Bin Khalifa University (HBKU; Approval No. QBRI-IRB 2021–03-078). Written informed consent was obtained from all study participants.

#### Demographic variables

In the MASC cohort, questionnaires were used to collect data on sex and age at each study site. Data were stored and managed using the Research Electronic Data Capture (REDCap) tool, a web-based electronic system hosted by the University of the Witwatersrand [23]. In the UKBB, data was collected at 22 assessment centres in the United Kingdom where they signed an electronic consent form. Self-completed electronic questionnaires and computer-assisted interviews were used to collect data ^49^. In QBB, all participants attended an assessment session in which physical measurements were collected, and each participant filled a standardized questionnaire reporting information on lifestyle, diet, and medical history ^48^.

#### Anthropometric measurements

A digital scale (model: TBF-410; Tanita Corporation, Arlington Heights, Illinois) was used to measure weight to the nearest 0.1kg, in people without shoes and with minimum clothing in MASC. Height was measured using a wall-fixed digital stadiometer (Holtain, Crymych, UK) ^13^. Weight, standing and sitting height were collected from participants in the UKBB and QBB.

#### Sample collection and analysis

Blood plasma was collected from MASC participants after a 10-12 hour overnight fast. OLINK Proteomics AB (Uppsala, Sweden) was used for proteomic analysis. Cardiovascular disease (CVD) panels II and III (www.olink.com) were used in the study. Performance of the proteomic assays was evaluated using four internal standards [30]. For UKBB participants, 9 ml of blood was collected from 54,219 participants, highly representative of the whole cohort, and fractionated into plasma, buffy coat, and red cells before storage at −80 °C prior to analysis. The antibody-based Olink Explore 3072 PEA was used for analysis ^38^. Normalized protein expression (NPX) values were used to report the results. A total of 180 proteins present in both the MASC and UKBB Biobank datasets were retained for analysis.

In the QBB cohort, serum glucose levels were measured using an enzymatic hexokinase method (GLUC3 kit, Roche, Switzerland) on a Cobas analyzer (Roche). Serum C-peptide and insulin concentrations were measured using sandwich electrochemiluminescence immunoassays (ECLIA) with Elecsys C-peptide and insulin kits (Roche) on the Cobas platform. Whole-blood HbA1c was measured using a turbidimetric inhibition immunoassay (TINIA) with the Tina-quant HbA1c Gen. 3 kit (Roche). Serum triglycerides were quantified using an enzymatic colorimetric method. Baseline levels of 1,305 circulating proteins were quantified in at Weill Cornell Medicine-Qatar using the SOMAscan aptamer-based proteomics platform (version 1.3; SomaLogic, Boulder, CO), as previously described ^50^. Raw intensity data were provided by SomaLogic following internal quality control and batch normalization procedures. Standard processing steps included hybridization normalization, median signal normalization, and signal calibration to adjust for inter-plate variation. No samples or data points were excluded from the study. Protein abundance values were log2-transformed and standardized before downstream analyses.

### BMI classification

Individuals were classified into measured BMI classes using the WHO cut-off as follows: underweight (BMI < 18.5 kg/m^2^), normal weight (BMI ≥ 18.5 and < 25kg/m^2^, overweight (BMI ≥ 25 and < 29.9 kg/m^2^ and obese (BMI ≥ 30 kg/m^2^).

### Proteomic score development

Proteomic scores were computed for South Africans of African Ancestry in MASC and four ancestral groups in the UKBB cohort: East Asians, South Asians, Africans, and Europeans. We applied, mean imputation for proteins exhibiting high missingness (>5%), specifically SPON1, BNP, TLT-2, NT-proBNP and CA5A, was performed to minimize bias and retain these biomarkers in our analyses. The protein biomarkers were log-transformed and scaled before analysis using adaptive LASSO with 10-fold cross-validation to select the optimal penalty. Model coefficients of the retained proteomic biomarkers were used to compute the weighted proteomic scores, which we denoted as MASC - contAFRPPS, East Asians - EASPPS, South Asians - SASPPS, Africans in the UKBB - diaAFRPPS, and Europeans - EURPPS. Cross-population generalizability was determined by i) assessing whether the ancestry BMI proteomic score was significant and had concordance direction of effect when it was applied across ancestry groups ii) computing out of sample mean trait BMI variance explained by each ancestry specific proteomic scores when it is applied to other ancestry groups excluding from the one it is derived from.

### Statistical Analysis

Analysis was done using R (version 4.4.0). Descriptive statistics were used to summarize baseline characteristics of the cohorts. Parametric tests were used to describe normally distributed continuous variables, and non-parametric tests were applied to those that were not normally distributed. Z-score standardization was used to transform the cardiometabolic traits and proteomics scores to enable stacked comparisons of associations on the same scale and in the same plot for better presentation in forest plots.

#### Cross-sectional generalizability and longitudinal prediction of BMI by the plasma proteomic scores

Linear regression models were used to assess the association between proteomic scores and BMI in multiple ancestries while adjusting for age and sex in the combined men and women datasets. The predictivity of proteomic scores was determined as the incremental variance (full model R² - null model R²). Mean out of sample variability in BMI explained by the proteomic scores was obtained by averaging the R^2^ of the cross-ancestry models, excluding the one in which the proteomic score was trained. The 95% confidence intervals were computed as mean ± 1.96 × SE. Generalizability was regarded as relatively similar mean outsample predictivity of the plasma proteomics, significant associations across ancestry with concordant effect direction.

For longitudinal prediction of BMI, only those without obesity at baseline were included in the analysis. We evaluated how baseline proteomic scores predicted incident obesity(**Figure 1A**). The median follow-up time between the two measurements was 6 years for QBB and 9 years for UKBB. Cox proportional hazards models, adjusted for age and sex, were used to predict the longitudinal risk of obesity using the ‘survival’ package of the R statistical software in the combined datasets of the UKBB and QBB. Stratified models were performed for men and women, adjusted for age. Schoenfield plots were used to check compliance with the PH assumptions.

We further determined the proteins that were common in the different scores from multiple ancestries. After determining the common proteins (eight proteins), we created a score by multiplying the ancestry specific LASSO model effect sizes with the protein values this was then used in our cross sectional and longitunal analysis.

#### Mendelian randomization

We performed two-sample MR analyses using BMI as the exposure and circulating protein concentrations as the outcomes. BMI genetic association data were obtained from the Million Veteran Program (MVP) GWAS, which included 424,221 participants of European ancestry ^36^. Protein outcome data were obtained from the UK Biobank Pharma Proteomics Project (UKB-PPP) proteomic GWAS, which quantified 2,496 proteins in 34,557 individuals of European ancestry using the Olink Explore 3072 platform ^37,38^. Genetic instruments were the leading variants identified in this study. Briefly, to identify statistically robust associations, we applied a genome-wide significance cutoff of 4.6×10^−1^^1^, adjusted according to the 1,038 effectively independent traits evaluated in the study. Independent association signals and their corresponding lead SNPs were subsequently extracted using linkage disequilibrium–based clumping implemented in PLINK 1.9 ^51^. This procedure followed a hierarchical two-step framework adapted from methods originally introduced in FUMA ^52^. The primary MR analysis used a random-effects inverse-variance weighted (IVW) approach implemented in TwoSampleMR v0.5.6 ^53^. A Bonferroni-corrected significance threshold of P < 6.3 × 10⁻³ (0.05/8) was applied. MR depends on three core assumptions: (1) the genetic instruments are associated with the exposure (relevance); (2) they are not linked to confounders of the instrument–outcome relationship (independence); and (3) they influence the outcome only through the exposure (exclusion restriction, or absence of horizontal pleiotropy). To mitigate weak-instrument bias, which threatens the relevance assumption in MR analysis, we calculated F-statistics and verified that they exceeded 10. We also conducted several sensitivity analyses to assess potential violations of the MR assumptions. Finally, MR analyses require genetic instruments that are strongly associated with the exposure and exert their effects primarily through the exposure of interest. Variants within the human leukocyte antigen (HLA) region are well known for extensive pleiotropy and complex linkage disequilibrium patterns ^54^, which may bias causal estimates by influencing outcomes through pathways unrelated to BMI. Therefore, consistent with established practice in GWAS-based MR studies of BMI, variants located within the HLA region (chromosome 6:25–34 Mb) were excluded from the analysis to minimize potential pleiotropic bias and improve the validity of the genetic instruments ^55^.

#### MR sensitivity analysis

We evaluated heterogeneity, directional horizontal pleiotropy (via the MR-Egger intercept), the MR-Egger slope estimate, and potential reverse causation. Heterogeneity was quantified using the *I*² statistic, with values >50% considered substantial. Directional pleiotropy was assessed using the MR-Egger intercept test, where P < 0.05 indicated evidence of pleiotropy. To obtain outlier-robust estimates, we conducted additional analyses using the weighted median and MR-Egger slope. Proteins had to demonstrate concordant directional effects across all approaches, including IVW, and were excluded if this criterion was not met. For reverse MR (testing whether plasma protein levels influence BMI) we used cis-pQTLs derived from UK Biobank. Depending on instrument availability, we applied either the IVW approach or the Wald ratio (for single-SNP instruments). Statistical significance was defined using a Bonferroni-corrected threshold.

#### Proteomic Prediction of Cardiometabolic Traits

To determine the utility of proteomic profiling in cardiometabolic risk prediction independent of BMI and waist circumference. We performed stratified analyses for the EURPPS, 8protEURPPS adjusted in separate models for BMI and waist circumference with age and sex as covariates for selected BMI-related cardiometabolic traits, namely VAT, subcutaneous fat, insulin sensitivity (represented by Matsuda Index), insulin resistance (represented by HOMA-IR), fasting glucose, 2-hour glucose, triglycerides, systolic blood pressure and diastolic blood pressure among MASC participants. Multiple testing correction was applied using the FDR correction.

### Recalibration with Elastic Net in MASC and QBB

To evaluate the discrepancies between the proteomic and measured BMI, we assessed their performance in the MASC and validated it in the QBB. The proteomic BMI scores in MASC and UK Biobank were derived from Olink proteomic measurements, whereas QBB proteomic data were generated using the SomaScan platform. Because coefficients derived from Olink are not directly transferable to SomaScan due to differences in assay technology, dynamic range, and signal scaling, the proteomic BMI model was recalibrated before downstream analyses. The analysis was restricted to proteins that overlapped between the MASC and UK Biobank cohorts.

Participants in MASC were randomly split into a training set (20%, n = 195) and testing set (80%, n = 756) while QBB participants were randomly split into a training set (30%; *n* = 725) and a testing set (70%; *n* = 1,685). An ElasticNet regression model was fitted to the training set with the measured BMI as the outcome and the eight common proteins (six in the QBB) as predictors. The coefficients obtained were used in the testing set to generate the proteomic score as the weighted sum of protein abundance. To place the score on the BMI scale, a linear regression model was fitted in the training set with measured BMI as the outcome and the raw proteomic BMI score as the predictor. The fitted model was then applied to the testing set to generate calibrated proteomic BMI estimates for the testing set.

### Discrepancy Between Proteomic BMI and Measured BMI

The discrepancy score was calculated as the difference between the proteomic BMI and measured BMI (pBMI − BMI). Positive values indicated that proteomic BMI exceeded measured BMI, whereas negative values indicated the opposite. Among individuals with normal-weight BMI (n = 188 in MASC and *n* = 461 in QBB), discrepancy quintiles were generated to identify individuals with a normal BMI but an overweight- or obesity-like proteomic profile. Age- and sex-adjusted marginal means of measured BMI and proteomic BMI were estimated across discrepancy quintiles.

### Discrepancy and Metabolic Trait Quintiles

To examine metabolic variation among normal-weight individuals, participants were stratified into quintiles of VAT, SAT, Triglycerides, Matsuda Index, HOMA-IR, QUICKI, the triglyceride-glucose (TyG) index and HDL cholesterol. Age- and sex-adjusted marginal means of measured BMI and proteomic BMI were estimated across metabolic trait quintiles. Linear trend tests were performed by fitting multivariable linear regression models with the metabolic trait quintile entered as an ordinal variable (coded 1–5) after adjusting for age and sex. The resulting regression coefficient (β) represented the linear trend across quintiles.

The proportion of variance in each metabolic trait explained by measured BMI and proteomic BMI was evaluated using linear regression models in the QBB. The null model included only age and sex. Separate models additionally included either measured BMI or proteomic BMI, whereas the full model simultaneously included age, sex, measured BMI, and proteomic BMI. The variance explained by each measure was calculated as the increase in adjusted *R*² relative to the null model (Δadjusted *R*²). The partially adjusted *R*², representing the unique variance explained by each BMI measure after accounting for the other, was calculated by comparing the full model with a reduced model excluding the BMI measure of interest. Confidence intervals for all *R*² estimates were obtained using 1,000 bootstrap resamples of the normal-weight validation set, with 95% percentile confidence intervals.

## Data availability

Proteomic and phenotype data used for this study are available upon request from the MASC investigators and UKBB data portal. The QBB data analyzed in this study can be obtained through an ISO-certified protocol. Requests to access these datasets should be directed to https://www.qphi.org.qa/research/how-to-apply.

## Code availability

All code used for data processing and statistical analyses are available upon request from the authors.

## Acknowledgements

The willing participation of study participants who provided the data and study staff who collected data for this study is sincerely acknowledged. This research has been conducted using the UK Biobank Resource under Application Number 20915. The MASC study is funded by the South African Medical Research Council with funds from the South African National Department of Health, Medical Research Council, UK (via the Newton Fund) and GSK Africa Non-Communicable Disease Open Lab (via a supporting Grant project Number: ES/N013891/1) and South African National Research Foundation (Grant no: UID:99108) for data collection in 2017/2018, and the European Research Area Personalised Medicine Joint Transnational Call (ERA-PerMed JTC2022), by the South African Medical Research Council (SAMRC) with funds received from the Department of Science and Innovation (data collection in 2023-2025). LJW is supported through a strategic grant [STRATGNT2023-01] from the DST-NRF Centre of Excellence in Human Development. LKM is supported by the South African Medical Research Council. Opinions expressed and conclusions arrived at are those of the authors and are not to be attributed to their institutions or funders. We thank the Qatar Precision Health Institute (QPHI) for providing the phenotypic data. Special thanks to all study participants for their invaluable contribution. YI is supported through the Canada Postdoctoral Research Award (CPRA) program from the Canadian Institute of Health (grant number 208352). *TC is supported by American Diabetes Association Grant* 1-26-ACE-0899.

## Author contributions

M.M. and T.C. conceptualized the study. M.M., T.C., Y.I., M.M., C.Z, R.N, U.A. and O.M.E.A. performed data processing and statistical analysis. M.M., T.C., Y.I., L.W., S.A.N., L.K.M, M.R., R.N., S.F., G.P., A.P.M. J.G., M.C., M.M., C.Z., P.W.F, A.B.K, U.A. and O.M.E.A. contributed to the writing, review and finalization of the manuscript.

